## Supplementary figures and images for "Selective whole-genome amplification reveals population genetics of *Leishmania braziliensis* directly from patient skin biopsies"

### Supplemental Figure 1

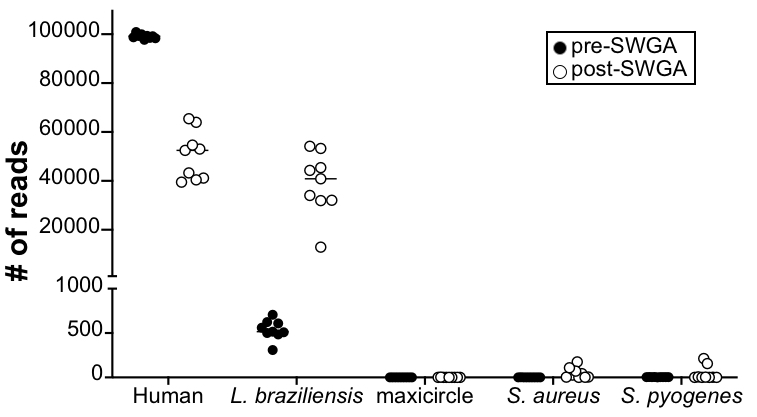

### Supplemental Figure 2

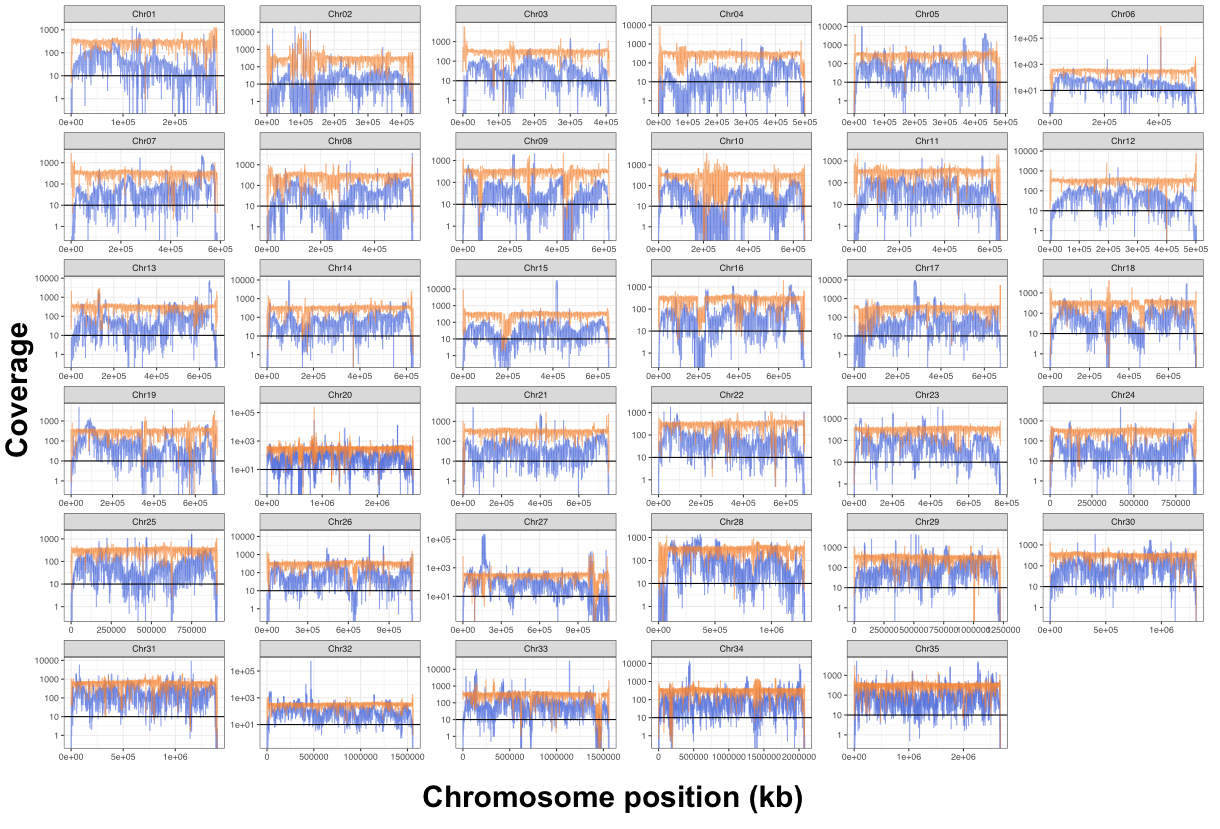
